# Conserved neuroectodermal aging encodes primate health and longevity

**DOI:** 10.64898/2026.05.05.26352498

**Authors:** Shaopeng Yang, Zhuoyao Xin, Wei Wang

## Abstract

Neuroectoderm-derived tissues are highly metabolically active and exhibit minimal regenerative turnover, rendering them uniquely vulnerable to age-related stress while preserving undiluted degenerative signals. Yet aging dynamics in these tissues remain elusive in living primates. Here, we introduce an *in vivo* neuroectodermal aging clock and trace its trajectory in 66,602 human adults and six rhesus macaques across nine health and disease cohorts using an *in situ* optical biopsy. Through a digital histology atlas integrated with artificial intelligence, we resolve tissue representations of neuroectodermal aging within the human retina, predominantly localized to the metabolically active ganglion and bipolar cell populations and the photoreceptor complex, while demonstrating their evolutionary conservation across primate species. Neuroectodermal aging predicts health and longevity, scales across space and time, and captures preclinical aging signals within and beyond the neuroectodermal compartment. This framework is further validated in a diabetic population, where robust prognostic and dynamic sensitivity are preserved across physiological and perturbed states. Our work establishes a scalable framework for resolving neuroectodermal aging in living primates and linking tissue-level vulnerability to systemic health trajectories.

## Introduction

Every vertebrate life begins with three germ layers established during gastrulation, an enduring blueprint that defines cellular lineages and their lifelong fates. Although development and aging appear as the bookends of life, evidence shows that aging programs are inscribed during embryogenesis and repurposed in adulthood to coordinate age-related stress.^1-3^ This framework supports distinct aging trajectories across lineages shaped by their divergent ontogenies. Neuroectodermal tissues mark the earliest sites of aging-program activation, where intense metabolic demand and intricate cytoarchitecture confer exceptional vulnerability to aging stress.^3-6^ Unlike endodermal and mesodermal derivatives that preserve regenerative plasticity to buffer degeneration, neuroectodermal tissues remain constrained by persistent suppression of post-developmental stemness.^7-10^ This absence of regenerative dilution of aging signals renders the neuroectoderm a purified model of intrinsic aging. Yet despite histological and cellular evidence of neuroectodermal aging traced back to embryonic patterning, its dynamics during primate adulthood remain uncharted.

While various approaches have been developed to quantify aging, referred to as aging clocks, they have thus far lacked *in vivo* histological definition.^11-15^ In contrast to upstream molecular clocks derived from blood sampling (e.g., DNA methylation, protein) that capture average drifts across heterogeneous organ chemistries, and downstream anatomical clocks (e.g., brain MRI, chest radiographs) that reflect consequences of organ deterioration, it remains largely unexplored how gradual molecular drifts consolidate into anatomical organ outcomes through mesoscale histological degeneration. Moreover, most molecular perturbations are transient and reversible, with inherently high plasticity allowing short-term physiological noise (e.g., diet, exercise) to obscure the slow yet irreversible signals of decline.^16,17^ This translates into a weak correspondence between molecular snapshots and underlying histological degeneration or anatomical outcomes. Conversely, macroscopic anatomical outcomes represent the opposite end of the spectrum, where accumulated degeneration no longer permits deconvolution to recover histological granularity.

The retina offers a natural window into neuroectodermal aging. As an evaginated domain of the brain that resides at the back of the eye, it is optically exposed through transparent ocular media, granting direct access to living neural tissue that elsewhere requires invasive or postmortem sampling.^18,19^ Unlike other intracranial brain regions (e.g., cerebral cortex), the retina retains a highly conserved blueprint across species, organized into three nuclear laminae for phototransduction and signal relay separated by two synaptic plexiform strata that channel signals in a defined direction.^20^ A further advantage over the cortex, where convoluted folding and interwoven axonal projections obscure histological boundaries, is that the retina preserves the planar, laminated organization of early neurogenesis, enabling direct mapping of cellular and subcellular domains at histological precision. Evidence shows that degeneration of the retinal tissues closely links to aging, with early manifestations detectable in multiple age-related diseases (ARDs) beyond the eye.^18,19,21-23^ Yet no study has systematically demonstrated how this laminated architecture supports aging assessment or validated it across diverse populations.

We hypothesize that neuroectodermal aging can be read out from the protruding, well-laminated retinal architecture. Leveraging a unique optical biopsy of the retina across eight large-scale health and disease cohorts, we resolve human neuroectodermal aging at real-time histological granularity and demonstrate its evolutionary conservation across primate species. Notably, application to a type 2 diabetes population allows evaluation of robustness across physiological and metabolically stressed states. Finally, we interrogate how localized vulnerabilities within the retinal laminae encode tissue signatures of neuroectodermal aging.

## Results

### A neuroectodermal aging clock from bilateral retinal sections resolving laminar tissue organization

To trace neuroectodermal aging from the planar, laminated architecture of the living human retina, we leveraged *in vivo* histological sections enabled by optical coherence tomography (OCT) in 66,602 diverse participants from eight cohorts (**Figs. 1a–b** and **Methods**). OCT serves as an optical biopsy by measuring the echo time delay and backscattered light intensity to provide retinal sections *in situ* and in real time.^24^ We established a digital histology atlas, CHRONOS, which integrates a fully automated preprocessing pipeline with a customized aging clock trained on bilateral retinal sections from 14,792 healthy individuals in the Chinese Ocular Imaging Project (COIP), the Guangzhou Twin Eye Study (GTES), and the Zhongshan Angle-closure Prevention (ZAP) project, to chart digital tissue representations underlying neuroectodermal aging (**Methods**).

**Fig. 1.**
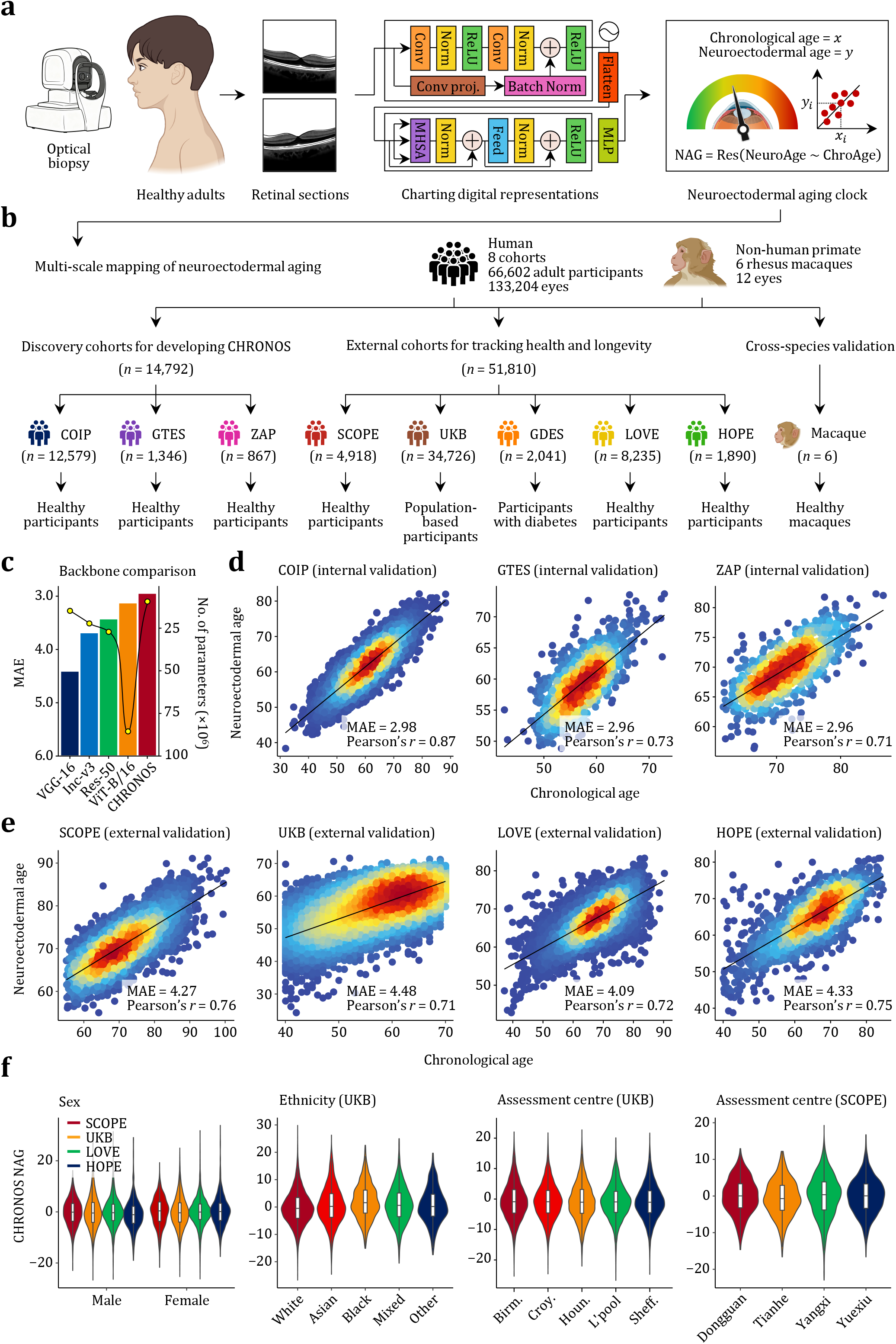
Development and validation of the neuroectodermal clock. **(a)**, Bilateral retinal sections from healthy adults acquired by *in vivo* optical biopsy were used to construct digital tissue representations of neuroectodermal aging. NAG was calculated as the residual from regressing neuroectodermal age on chronological age. **(b)**, Multi-scale mapping of neuroectodermal aging across nine human and macaque cohorts. The COIP, GTES and ZAP cohorts of healthy adults were used to develop CHRONOS (n = 14,792), whereas the SCOPE, UKB, GDES, LOVE and HOPE cohorts were used for external validation and for tracking health and longevity (*n* = 51,810). Six healthy rhesus macaques were used for cross-species validation. (**c**), Backbone comparison of CHRONOS performance and computational efficiency relative to alternative deep-learning architectures (*n* = 14,792). Bars indicate mean absolute error, and the line denotes numbers of parameters used. (**d**), Performance of the neuroectodermal aging model in three internal healthy cohorts (COIP *n* = 12,579; GTES n = 1,346; ZAP *n* = 867) evaluated using five-fold nested cross-validation on holdout test sets. (**e**), Performance in three external healthy cohorts (SCOPE *n* = 4,918; LOVE *n* = 8,235; HOPE **n** = 1,890) and an external population-based cohort (UKB *n* = 34,726). (**f**), First panel: sex distributions of NAG in the SCOPE (female *n*⍰=l2,153; male *n*⍰=l2,765), UKB (female *n*⍰=l18,525; male *n*⍰=l16,201), LOVE (female *n*⍰=l4,649; male *n*⍰=l3,586), and HOPE (female *n*⍰=l1,264; male *n*⍰=l626). Second panel: distributions of NAG according to self-reported ethnicities in the UKB (White *n*⍰=l31,383; Black *n*⍰=l1,068; Asian *n*⍰=l1,205; other *n*⍰=l232; mixed *n*⍰=l838). Third and fourth panels: distributions of NAG according to assessment centres in the UKB (Birmingham *n*⍰=l7,972; Croydon *n*⍰=l8,772; Hounslow *n*⍰=l6,620; Liverpool *n*⍰=l2,579; Sheffield *n*⍰=l8,675) and SCOPE (Dongguan *n*⍰=l387; Tianhe *n*⍰=l608; Yangxi *n*⍰=l2,907; Yuexiu *n*⍰=l1,016). Overlaid boxplots indicate the median (centre line) and interquartile range (box bounds), with whiskers extending to the minimum and maximum within 1.5× the interquartile range. Birm. = Birmingham; Croy. = Croydon; Houn. = Hounslow; L’pool = Liverpool; Sheff. = Sheffield.

To assess whether tissue representations within the neuroectodermal retina are generalizable across populations, we applied CHRONOS to 51,810 transethnic participants from five independent health and disease cohorts (**Fig. 1b** and **Methods**). These included three healthy cohorts: the High-definition Oculo-Phenomics Evaluation (HOPE) study, the Landscape of Optic Neuropathy and Vasculopathy Evolution (LOVE) study, and the Southern China Ophthalmic Epidemiology (SCOPE) study; a population-based cohort with mixed health status, the UK Biobank (UKB); and a diabetes cohort, the Guangzhou Diabetic Eye Study (GDES). Baseline characteristics of all cohorts are summarized in **Supplementary Table S1**.

The CHRONOS clock is a convolutional Transformer neural network optimized after comparative evaluation of architecture performance and efficiency (**Fig. 1c** and **Methods**). This hybrid architecture leverages convolutional modules to extract fine-grained laminar tissue textures within the retina, while the Transformer encoder integrates long-range dependencies to perform contextual reasoning across its laminated organization. Across healthy populations, CHRONOS faithfully recapitulated the chronological aging trajectory both in internal and external population (**Figs. 1d–e**).

We and others have previously shown that the gap between predicted and chronological age serves as a measure of an individual’s biological age relative to their same-aged peers.^11-15^ Based on this concept, we defined the neuroectodermal aging gap (NAG) as the residual from regressing CHRONOS age on chronological age, reflecting tissue aging signals within the neuroectodermal retina unaccounted for by chronological age (**Methods**). NAG distributions were comparable across cohorts by sex, across self-reported ethnicities in the UKB, and across the nine assessment centres in both the UKB and SCOPE (**Fig. 1f**).

### Neuroectodermal aging is a strong predictor of cellular aging hallmarks and mortality risk

If CHRONOS reflects fundamental processes of intrinsic aging, it should be linked to one or more of the original hallmarks of cellular aging.^25,26^ To determine this, we examined the associations of NAG with the prevalence and incidence of the nine aging hallmarks: genomic instability (GI), telomere attrition (TA), epigenetic alterations (EA), loss of proteostasis (LP), deregulated nutrient sensing (DNS), mitochondrial dysfunction (MD), cellular senescence (CS), stem cell exhaustion (SCE), and altered intercellular communication (AIC) (**Fig. 2a** and **Methods**). These hallmarks were operationalized using 83 ARDs validated to represent aging hallmarks through a multistep framework^27^ integrating human aging corpus mining, shared genetic signals from the genome-wide association study (GWAS) catalogue, gene set enrichment analysis, and disease co-occurrence networks (**Supplementary Table S2**).

**Fig. 2.**
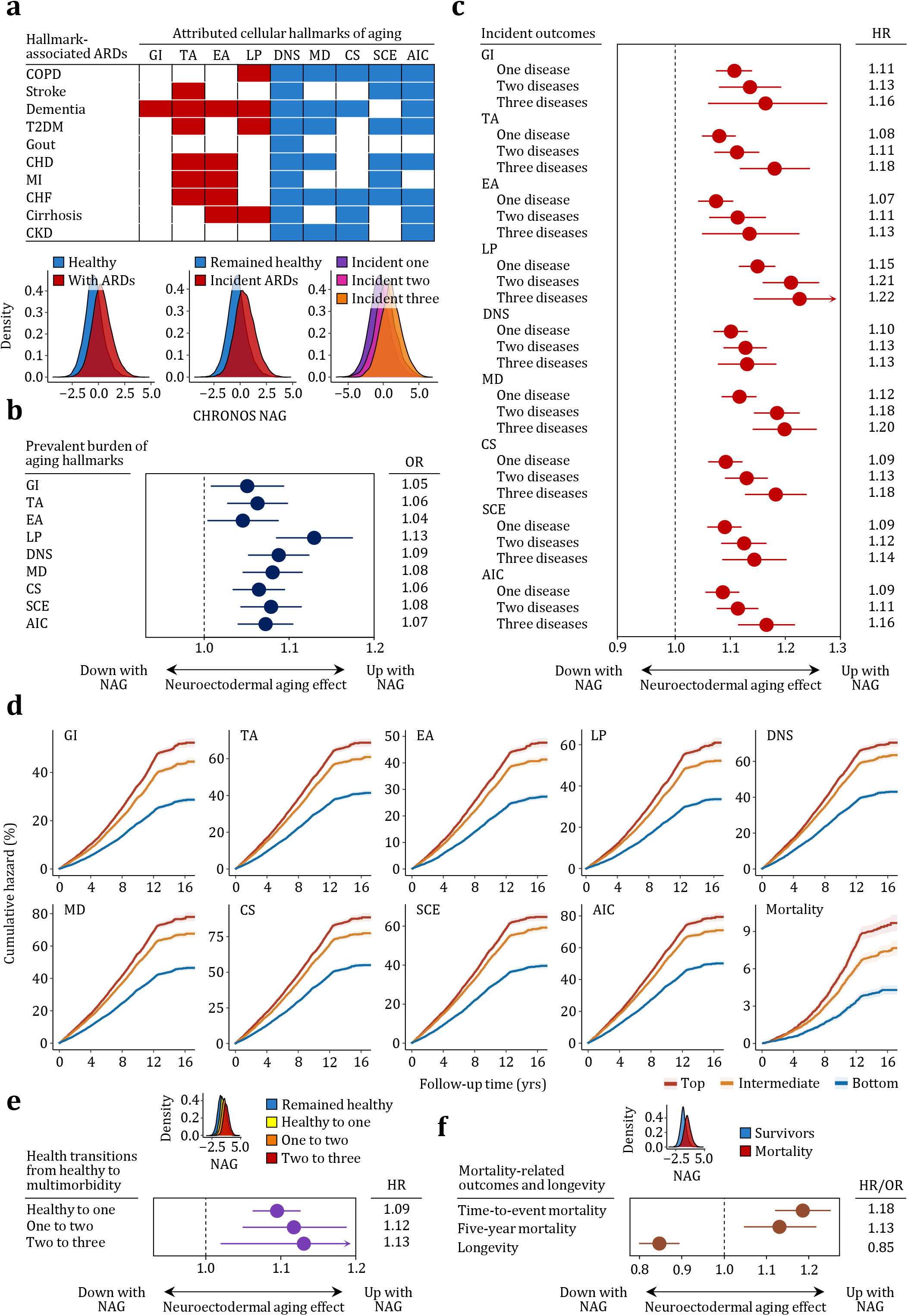
Neuroectodermal aging is a strong predictor of cellular aging hallmarks and mortality risk. (**a**), Mapping of hallmark-associated ARDs to their attributed original cellular hallmarks of aging. A full list of ARDs is shown in Supplementary Table S2. Density distributions show NAG in baseline healthy individuals (*n* = 23,777) or those with a diagnosis of any of the 83 ARDs (*n* = 10,936), baseline healthy individuals who remained healthy (*n* = 11,127) or developed incident ARDs during 16 years of follow-up (*n* = 12,650), and those who developed one (*n* = 4,420), two (*n* = 2,945) or more than three (*n* = 5,285) incident ARDs in the UKB. (**b**), Associations of NAG with prevalent burden of hallmark-specific ARDs, estimated using multivariable logistic models (*n* = 34,713). Circles represent the estimated ORs, with 95% CIs indicated as lines of error bars. All ORs remain statistically significant through two-sided Wald tests after controlling FDR for multiple testing. (**c**), Associations of NAG with incident hallmark-specific ARDs during follow-up period, estimated using multivariable CPH models (*n* = 23,777). Effect estimates and 95% CIs are shown as in b. All HRs remain statistically significant through two-sided Wald tests after controlling FDR for multiple testing. (**d**), Cumulative hazard throughout the follow-up period for developing hallmark-specific ARDs and mortality, stratified by NAG tertiles (*n* = 23,777). Data are presented as observed event rates with 95% CIs shown as shading derived from survival proportions. (**e**), Associations of NAG with health transitions from a disease-free baseline to ARD multimorbidity during follow-up period, estimated using multivariable CPH models (*n* = 23,777). Effect estimates and 95% CIs are shown as in b. All HRs remain statistically significant through two-sided Wald tests after controlling FDR for multiple testing. (**f**), Associations of NAG with mortality and longevity, estimated using multivariable CPH or logistic models as appropriate (*n* = 23,777). Effect estimates and 95% CIs are shown as in b. All HRs and ORs remain statistically significant through two-sided Wald tests after controlling FDR for multiple testing. ARD = age-related disease; COPD = chronic obstructive pulmonary disease; T2DM = type 2 diabetes mellitus; CHD = coronary heart disease; MI = myocardial infarction; CHF = congestive heart failure; CKD = chronic kidney disease; GI = genomic instability; TA = telomere attrition; EA = epigenetic alterations; LP = loss of proteostasis; DNS = deregulated nutrient sensing; MD = mitochondrial dysfunction; CS = cellular senescence; SCE = stem cell exhaustion; AIC = altered intercellular communication; OR = odds ratio; FDR = false discovery rate; HR = hazard ratio.

Advanced NAG was strongly associated with the burden of prevalent ARDs across all nine hallmarks after adjustment for chronological age, sex, and major sociodemographic and lifestyle confounders (**Fig. 2b** and **Methods**). The strongest association was observed for LP, highlighting protein homeostasis as a key process underlying neuroectodermal aging and motivating preliminary proteomic profiling of NAG (**Supplementary Table S3**). In addition, all associations with the other hallmarks remained robust following multiple testing correction and were generally stronger for compensatory and integrative hallmarks (DNS, MD, CS, SCE, AIC) than for most primary hallmarks (GI, TA, EA). These findings suggest that neuroectodermal aging converges with broader systemic aging processes that coordinate cellular homeostasis and intercellular signalling rather than remaining confined to its local tissue context.

We next assessed whether neuroectodermal aging served as a predictor of future ARD risk (**Methods**). Leveraging >16 years of follow-up in healthy UKB participants, advanced NAG conferred higher risks of incident ARDs across all nine aging hallmarks in time-to-event analyses using multivariable Cox proportional hazards models (**Fig. 2c**). Within each hallmark, hazard ratios increased progressively with the incidence of one, two, and three ARDs, indicating a dose-response relationship between neuroectodermal aging and susceptibility to hallmark-related multimorbidity. Stratification by NAG deciles further revealed sharply divergent trajectories of cumulative incidence, with significantly higher incident rates among individuals in the top decile compared to those in the middle or bottom (**Fig. 2d**).

To disentangle health transitions separately, we performed stepwise analyses of ARD multimorbidity development from a disease-free baseline (**Methods**). Advanced NAG was associated with higher risks at each transition after multiple testing correction, including from zero to one, one to two, and two to three ARDs, with escalating hazards as baseline multimorbidity increased (**Fig. 2e**). These findings suggest that neuroectodermal aging not only tracks the initial onset of individual ARDs but also parallels the accelerated accumulation of ARD multimorbidity, reflecting the progressive loss of homeostatic resilience across neuroectodermal aging.

We further evaluated mortality and longevity, the terminal manifestations of aging, across several distinct definitions: (1) all-cause mortality assessed with time-to-event analysis; (2) short-term mortality in five years; and (3) longevity defined as survival beyond age 80 (**Methods**). Each SD advanced in NAG conferred a 12% higher risk (95% CI: 1.06 to 1.18) of all-cause mortality after multivariable adjustment (**Fig. 2f**), an effect size exceeding that of epigenetic and most established organ clocks trained on chronological age.^28,29^ Significant associations of attenuated NAG were also observed with both short-term mortality and longevity (**Fig. 2f**). Stratification by NAG deciles revealed divergent survival trajectories, with the highest risks observed among individuals in the top decile (**Fig. 2d**).

To test whether elevated mortality introduced competing risk bias into incident hallmark-related findings, we performed competing risk analyses (**Methods**). All associations between NAG and hallmark-related ARDs remained robust, indicating that mortality did not confound the observed results (**Supplementary Table S4**). Collectively, these findings demonstrate that neuroectodermal aging mirrors core hallmarks of organismal aging and provides a robust, biologically grounded readout of health and longevity.

### Neuroectodermal aging reveals frailty and systemic aging signals preceding clinically manifest conditions

Given the broad and robust associations of neuroectodermal aging with hallmark-related aging outcomes, we next asked whether it reflects physiological aging signals that precede clinically manifest conditions. Associations of NAG were evaluated with three complementary dimensions of aging physiology: (1) a comprehensive frailty index; (2) 15 individual biological aging markers; and (3) 16 individual physical and cognitive aging markers (**Methods**).

After multivariable adjustment and multiple testing correction, advanced NAG was significantly associated with most aging markers (**Fig. 3**). The frailty index ranked among the strongest associations, underscoring the ability of neuroectodermal aging to capture global vulnerability and homeostatic decline. Among biological aging signals (**Fig. 3a**), NAG was associated with renal function (cystatin C, creatinine), hepatic function (alanine aminotransferase [ALT], γ-glutamyl transferase [GGT], albumin), lipid metabolism (total cholesterol, triglycerides, LDL-c, HDL-c), and other landmark aging markers including telomere length, insulin-like growth factor 1 (IGF-1), HbA1c, and C-reactive protein (CRP). Among physical and cognitive markers (**Fig. 3b**), associations were observed with anthropometry (body mass index [BMI], waist-hip ratio), cardiovascular markers (systolic blood pressure [SBP], diastolic blood pressure [DBP], arterial stiffness index), sleep (sleeping ≥10 hours/day, insomnia), perceived aging markers (self-rated health, perceived facial aging, tiredness/lethargy), neurocognition (reaction time, fluid intelligence score), and lung function.

**Fig. 3.**
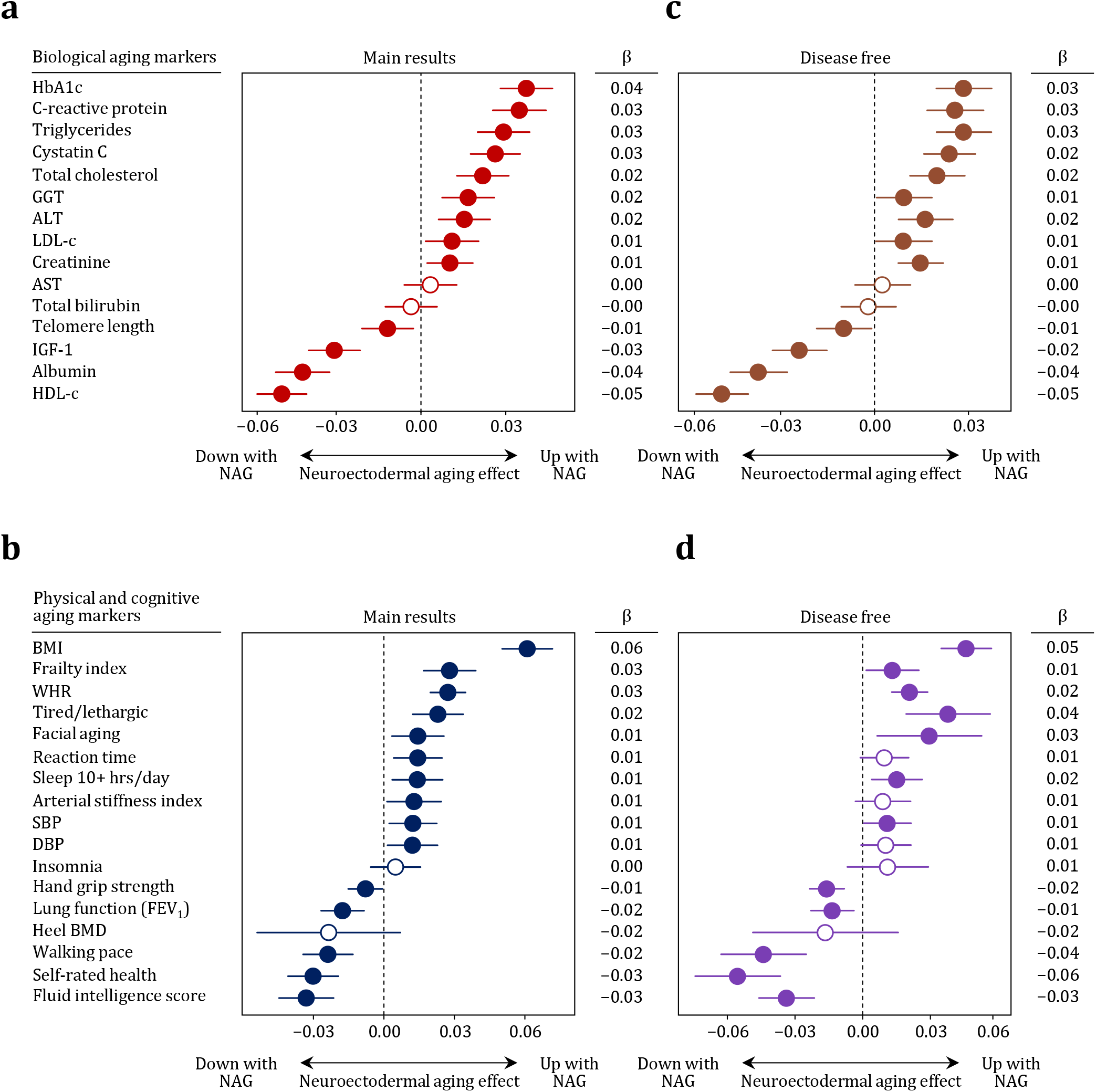
Neuroectodermal aging reveals frailty and systemic aging signals preceding clinically manifest conditions. **(a–b)**, Associations of NAG with biological, physical, and cognitive aging markers in the UKB, estimated using multivariable linear or logistic models as appropriate (*n* = 34,726). Circles represent estimated coefficients, with 95% CIs indicated as lines of error bars. *P* values were calculated through two-sided Student’s *t* or Wald tests after controlling FDR for multiple testing. Solid circles indicate statistical significance. (**c–d**), As in **a–b**, but restricted to individuals without any of the 83 age-related diseases studied (*n* = 34,726). Effect estimates and 95% CIs are shown as in **a–b**. HbA1c = hemoglobin A1c; GGT = γ-glutamyl transferase; ALT = alanine aminotransferase; LDL-c = low-density lipoprotein cholesterol; AST = aspartate aminotransferase; IGF-1 = insulin-like growth factor 1; HDL-c = high-density lipoprotein cholesterol; BMI = body mass index; WHR = waist-hip ratio; SBP = systolic blood pressure; DBP = diastolic blood pressure; FEV_1_ = forced expiratory volume in one second; BMD = bone mineral density.

To test whether these associations could be biased by reverse causation from underlying disease, we repeated analyses in individuals free of all 83 ARDs at baseline (**Figs. 3c–d**). In this disease-free subset, 28 of 32 associations persisted, indicating that NAG reflects intrinsic aging processes rather than pre-existing pathology. Persistent associations included telomere length, IGF-1, HbA1c, CRP, all renal, hepatic, and lipid markers, anthropometry, perceived aging, neurocognition, and lung function. Only three cardiovascular markers (SBP, DBP, arterial stiffness index) and insomnia lost significance after multiple testing correction.

Together, these findings establish neuroectodermal aging as a robust integrative marker of systemic frailty and physiological decline, capturing intrinsic aging processes beyond clinically manifest ARD outcomes.

### Conservation across health and diabetes, temporal and spatial scales, and primate species

To extend CHRONOS to metabolically perturbed contexts, we applied it to the GDES, a prospective diabetes cohort with biennial retinal tomography and up to four repeated visits (**Methods**). At baseline visit (*t*_0_), individuals with type 2 diabetes exhibited greater deviations between their neuroectodermal and chronological age (MAE = 5.21, Pearson’s *r* = 0.43) compared to healthy individuals (**Fig. 4a**). Each SD advanced in NAG was associated with 19% higher odds (95% CI: 12% to 26%) of baseline multimorbidity after multivariable adjustment, supporting generalized neuroectodermal aging in prevalent ARDs. Leveraging longitudinal follow-up for nine ARDs, advanced NAG was a strong predictor of incident ARDs spanning all cellular aging hallmarks (**Fig. 4b** and **Supplementary Table S2**). Associations were further replicated with physiological aging markers in the diabetic population, including frailty, renal function, lipid metabolism, HbA1c, CRP, anthropometry, SBP, sleep, perceived aging, and lung function (**Fig. 4b**).

**Fig. 4.**
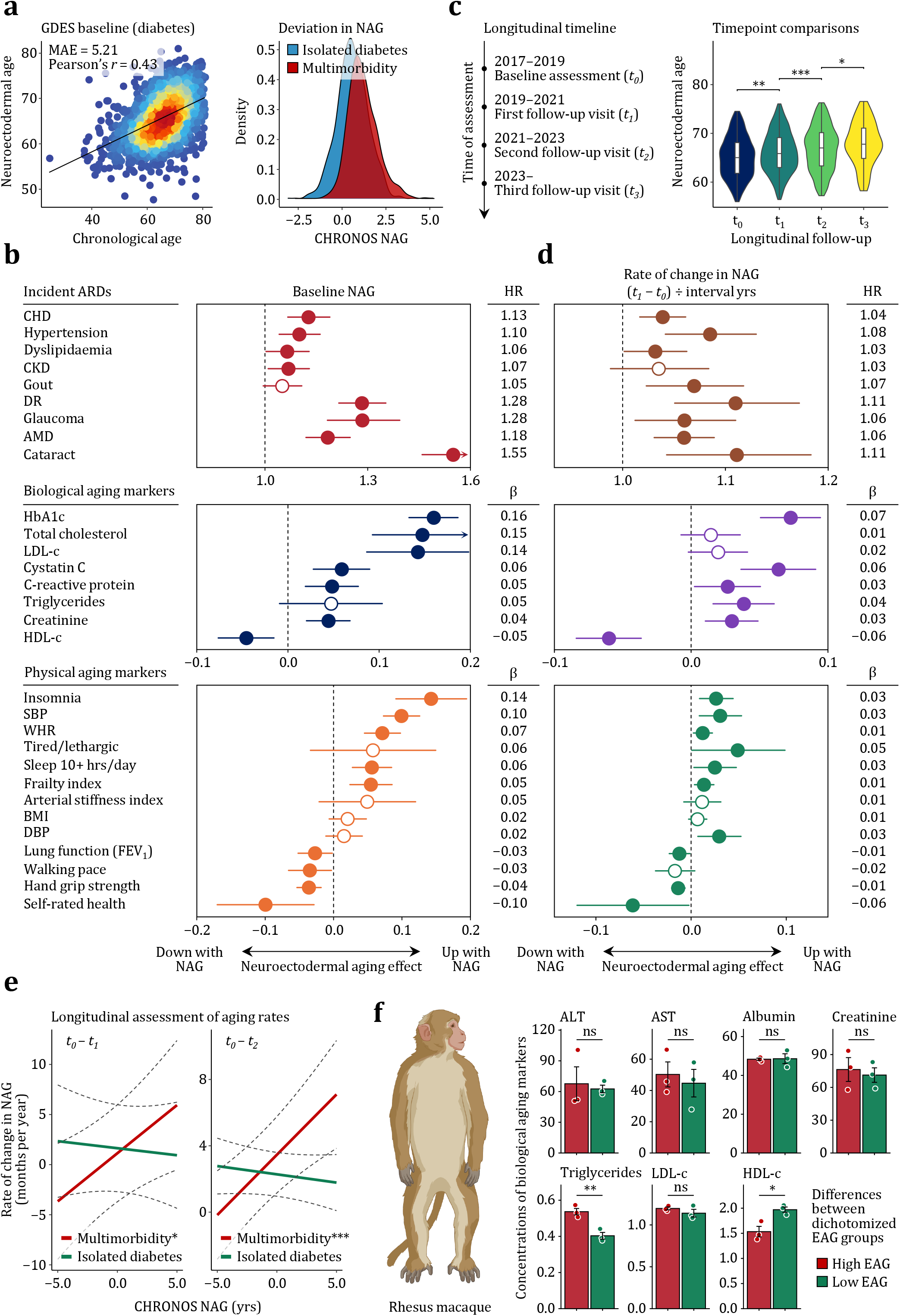
Contextual and evolutionary conservation of neuroectodermal aging. (**a**), Performance of the neuroectodermal aging model in the GDES. Density distributions show NAG in individuals with isolated diabetes (*n* = 1,145) or with multimorbidity (*n* = 896) at baseline. (**b**), Associations of NAG with incident ARDs and biological, physical, and cognitive aging markers, estimated using multivariable CPH, linear, or logistic models as appropriate (*n* = 2,041). Circles represent estimated coefficients, with 95% CIs indicated as lines of error bars. *P* values were calculated through two-sided Wald or Student’s *t* tests after controlling FDR for multiple testing. Solid circles indicate statistical significance. (**c**), Assessment timeline and longitudinal comparisons of neuroectodermal aging across four timepoints (*t*_0_ *n* = 2,041; t_1_ *n* = 1,352; t_2_ *n* = 1,533; t_3_ *n* = 720). Violin plots show NAG distributions, with overlaid boxplots indicating the median (centre line) and interquartile range (box bounds). Pairwise comparisons between adjacent timepoints were estimated using a linear mixed-effects model with individual-specific random intercepts. *P* values with Tukey adjustment were derived from two-sided Wald tests (t_0_ vs. t_1_, *P* = 0.003; t_1_ vs. t_2_, *P* = 6.20×10^−5^; t_2_ vs. t_3_, *P* = 0.015). Linear trends across time were assessed by modelling time as a continuous fixed effect in the same mixed-effects framework (*P* for trend = 4.2×10^−20^). Asterisks indicate statistical significance (\*\*\**P*⍰<0.001, \*\**P*⍰<0.01, \**P*⍰<0.05). (**d**), As in **b**, but for associations of the rate of change in NAG with incident ARDs and biological, physical and cognitive aging markers (*n* = 1,125). Effect estimates and 95% CIs are shown as in **b**. (**e**), Faster neuroectodermal aging among individuals with multimorbidity compared with those with isolated diabetes between t_0_–t_1_ (*n* = 1,125) and t_0_–t_2_ (*n* = 1,209). Lines of best fit show associations between the rate of change in EAG and the average age gap across the two visits, with 95% CIs indicated as dashed lines. Associations were tested separately in individuals with multimorbidity and those with isolated diabetes using two-sided Student’s t tests (t_0_–t_1_, *P*_*multimorbidity*_ = 0.012; t_0_–t_2_, *P*_*multimorbidity*_ = 1.74×10^−4^). Asterisks are defined as in **c**. (**f**), Cross-species validation in rhesus macaques (*n* = 6). Bars represent mean values, with 95% CIs indicated as error bars. Differences in biological aging markers between dichotomized NAG groups were assessed using two-sided Wilcoxon rank-sum tests (Triglycerides, *P* = 0.006; HDL-c, *P* = 0.034). Asterisks are defined as in **c**. MAE = mean absolute error; CHD = coronary heart disease; CKD = chronic kidney disease; DR = diabetic retinopathy; AMD = age-related macular degeneration; HbA1c = hemoglobin A1c; LDL-c = low-density lipoprotein cholesterol; HDL-c = high-density lipoprotein cholesterol; SBP = systolic blood pressure; WHR = waist-hip ratio; BMI = body mass index; DBP = diastolic blood pressure; FEV_1_ = forced expiratory volume in one second.

To track within-individual trajectories over time, we analysed four consecutive visits in the GDES (**Methods**). We observed across all timepoints that applying CHRONOS yielded stable performance (baseline [*t*_0_]: mean absolute error (MAE) = 5.21, Pearson’s *r* = 0.43; first follow-up [*t*_1_]: MAE = 5.35, Pearson’s *r* = 0.42; second follow-up [*t*_2_]: MAE = 5.29, Pearson’s *r* = 0.49; third follow-up [*t*_3_]: MAE = 5.39, Pearson’s *r* = 0.42), indicating consistent estimation under sustained metabolic stress. Intriguingly, neuroectodermal age showed a stepwise, monotonic progression with chronological time, as confirmed by pairwise comparisons between adjacent timepoints and trend analyses across the series (**Fig. 4c**). This trajectory aligns closely with the expected course of physiological aging over time, with observed stable variance suggesting that the apparent progression was unlikely to reflect measurement noise or model drift.

The repeated assessments enabled estimation of longitudinal rates of change in individual neuroectodermal aging (**Methods**). We focused primarily on NAG change between baseline (*t*_0_) and the first follow-up (*t*_1_), which provided the largest sample size while preserving sufficient follow-up time for incident outcome assessment. Strikingly, the rate of NAG change itself was a strong predictor of incident ARDs, independent of baseline NAG, chronological age, and the major confounders. (**Fig. 4d** and **Methods**). As with baseline NAG, the associations with the rate of NAG change extended to frailty, six of eight biological aging signals tested, and nine of 12 physical aging signals after multivariable adjustment (**Fig. 4d**). All associations remained robust after multiple testing correction, except for sleep duration and walking pace.

Among individuals with diabetes complicated by multimorbidity, we observed a clear coupling between average NAG over two timepoints and its longitudinal progression across follow-up intervals (**Fig. 4e** and **Methods**). From baseline (*t*_0_) to the first follow-up (*t*_1_), each additional year increase in average NAG corresponded to a 0.96-month/year (95% CI: 0.21 to 1.71) faster rate of neuroectodermal aging. A similar pattern was reproduced between baseline (*t*_0_) and the second follow-up (*t*_2_), with an increase of 0.73-month/year (95% CI: 0.35 to 1.10) in the rate of neuroectodermal aging for each year advanced in NAG. In contrast, individuals with isolated diabetes exhibited a near-constant rate of neuroectodermal aging over time, suggesting that accelerated neuroectodermal aging is amplified driven by ARD comorbidity.

We next queried whether CHRONOS traces conserved neuroectodermal aging beyond humans by applying it to six rhesus macaques (**Methods**). Although derived entirely from humans, neuroectodermal age exhibited a monotonic increase with chronological macaque age without any calibration (Spearman’s ρ = 0.89, 95% CI: 0.60 to 0.98), supporting conserved aging trajectories across species. After rescaling ages to the macaque lifespan using sexual maturity and maximum lifespan anchors (**Methods**), CHRONOS produced macaque estimates with an MAE of 2.1 years and a Pearson’s *r* of 0.74 against chronological age. Although the small sample size limits statistical power, macaques with advanced cross-species NAG exhibited significantly higher triglyceride and lower HDL-c levels than those with attenuated NAG (**Fig. 4g**). Consistent trends were observed for hepatic and renal function where advanced NAG accompanied apparent elevations in transaminases and creatinine.

### Interpretability of CHRONOS resolves tissue vulnerabilities of neuroectodermal aging

The interpretability of CHRONOS is pivotal for its biological and clinical translation. To interrogate the internal mechanisms by which CHRONOS encodes neuroectodermal aging, we developed a robust pipeline to map digital representations onto retinal tissues most vulnerable to the aging process across healthy and diabetic populations (**Fig. 5a** and **Methods**). Uniform manifold approximation and projection (UMAP) of CHRONOS embeddings revealed a smooth continuum across chronological age among healthy individuals in both internal and external population (**Fig. 5b**), aligning with a temporally ordered aging trajectory rather than stochastic variation. In contrast, this continuum was attenuated in the diabetic population, as would be expected for perturbed aging in ARDs.

**Fig. 5.**
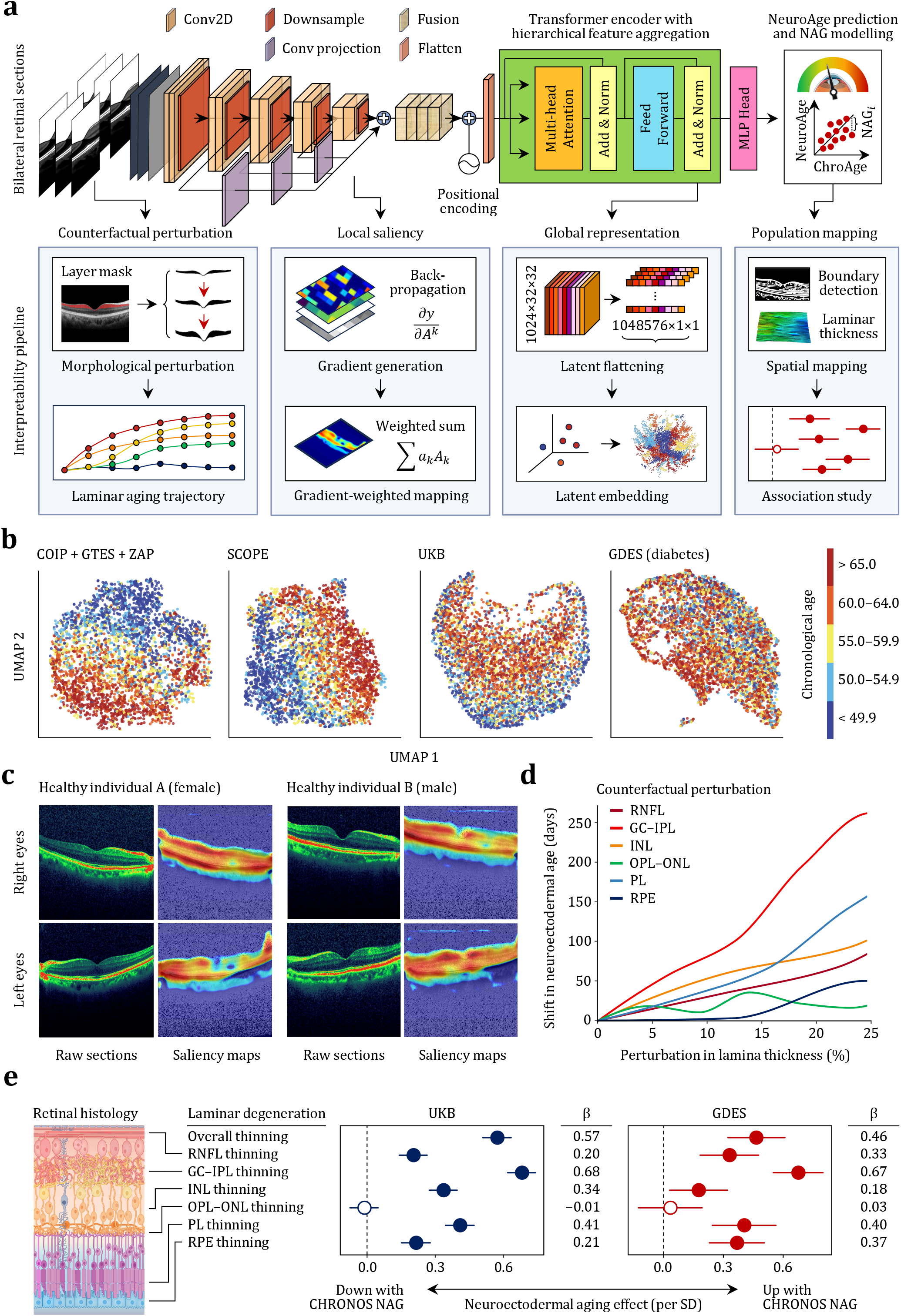
Interpretability of CHRONOS resolves tissue vulnerabilities of neuroectodermal aging. (**a**), Overview of the interpretability pipeline. (**b**), UMAP visualization of CHRONOS embeddings in internal (COIP⍰+ ⍰GTES⍰+ ⍰ZAP, *n* = 14,792) and external population (SCOPE *n* = 4,918; UKB *n* = 34,726; GDES *n* = 2,041). Each cohort was randomly downsampled to a maximum of 5,000 datapoints for visualization. (**c**), Gradient-weighted activation maps for representative bilateral retinal sections from a healthy female (left) and a healthy male (right) individual. (**d**), Shifts in neuroectodermal age estimates obtained through counterfactual simulations in which controlled morphological perturbations were introduced to individual retinal lamina (*n* = 1,000). (e), Associations between NAG and laminar degeneration across the three-dimensional retinal architecture in the UKB (*n* = 34,726) and GDES (*n* = 2,041), estimated using multivariable linear models. Circles represent estimated coefficients, with 95% CIs indicated as lines of error bars. *P* values were calculated through two-sided Student’s t tests after controlling FDR for multiple testing. Solid circles indicate statistical significance. RNFL = retinal nerve fibre layer; GC–IPL = ganglion cell–inner plexiform complex layer; INL = inner nuclear layer; OPL–ONL = outer plexiform layer–outer nuclear layer; PL = photoreceptor layer; RPE = retinal pigment epithelium.

Gradient-weighted activation mapping enabled refined spatial localization of structural vulnerabilities that contributed most strongly to the neuroectodermal aging process (**Methods**). The resulting aging saliencies were concentrated within the neuroectodermal retina rather than in deeper ocular tissues (e.g., the mesoderm-derived choroid), reflecting optical attenuation below the melanin-rich pigment epithelium that naturally confines sensitivity to the neuroectodermal retina. Within the retina, we identified a bimodal pattern of focal contributions concentrated in both inner and outer laminae, whereas intermediate lamina contributed minimally (**Fig. 5c**). These patterns parallel clinical and histological observations that certain retinal sublayers are disproportionately vulnerable to metabolic stress and implicated in multiple ARDs.^22,30-33^

As the retina preserves the planar and laminated blueprint of early neurogenesis, the spatially resolved saliency can be mapped onto discrete cellular and subcellular domains (**Figs. 5c** and **e**). Inner retinal signals primarily arose from ganglion and bipolar cell populations, which constitute the intermediate relay of the visual pathway.^34^ In tomographic sections, these correspond to the retinal nerve fibre layer (RNFL), formed by bundled ganglion-cell axons, and to the ganglion cell–inner plexiform complex layer (GC–IPL) and inner nuclear layer (INL), which house ganglion- and bipolar-cell somata and the synapse-rich dendritic circuitry connecting them.^19^ Outer retinal signals localized to the photoreceptor–RPE complex responsible for phototransduction, corresponding to the photoreceptor layer (PL) containing rod and cone segments and RPE layer composed of pigmented epithelial cells.^34^

To further probe biological and causal relevance, we conducted counterfactual simulations by introducing controlled morphological perturbations to individual retinal lamina (**Methods**). Although laminar degeneration is not necessarily symmetrical, this manipulation recapitulates the thickness variations typically reported in the aging retina. Perturbing laminar architecture induced distinct shifts in neuroectodermal aging estimates, suggesting that structural variation in the retinal laminae shapes the aging signals captured by CHRONOS (**Fig. 5d**). The most pronounced effects were identified in the GC–IPL, INL, and PL, where a 10% perturbation in thickness corresponded to 80-, 77-, and 51-day shifts in neuroectodermal aging. The RNFL and RPE exerted subtler yet monotonic influences, while the effects of the intermediate lamina fluctuated around null, suggesting structural resilience to aging trajectories.

To extend these findings from local sections to the spatial organization, we reconstructed the three-dimensional laminar architecture from consecutive sections and quantified laminar thickness across the neuroectodermal retinal landscape using a validated graph search approach based on multiscale gradient information across both healthy and diabetic populations (**Methods**). In the UKB, the average retinal thickness was strongly associated with NAG, with the GC–IPL, PL, and INL again showing the most pronounced associations after multivariable adjustment and multiple testing correction (**Fig. 5e**), concordant with laminae identified by gradient-weighted mapping and counterfactual perturbating. The RNFL and RPE showed weaker yet robust associations, whereas the intermediate lamina remained nonsignificant. All laminar associations with NAG were reproduced in the diabetic population (**Fig. 5e**).

## Discussion

This study establishes a translational framework for real-time, *in situ* profiling of neuroectodermal aging at histological granularity in adult organisms. By interrogating the retina’s uniquely laminated tissue through CHRONOS, we demonstrate that neuroectodermal aging reveals systemic health, predicts mortality and longevity, and captures preclinical aging signals within and beyond the neuroectodermal compartment. This approach is entirely non-invasive, conserved across spatial and temporal scales, and thus bridges microscopic tissue degeneration with macroscopic aging trajectories. Tissue-level mapping localized neuroectodermal aging predominantly to the metabolically active ganglion and bipolar cell populations and the photoreceptor complex, an organization that is evolutionarily conserved and recapitulated in rhesus macaques. Finally, application to a diabetic population reproduces the prognostic and dynamic sensitivity under sustained metabolic stress, underscoring the context-robust capacity of CHRONOS to operate across physiological and perturbed states as a unified framework for longitudinal health surveillance.

Organ-specific aging clocks can be structured into three hierarchical tiers that capture the molecular, histological, and anatomical manifestations of aging. Leveraging retina’s unique accessibility, we present the first *in vivo* aging clock operating at mesoscale histological resolution, providing intermediate stepping stones between transient molecular drifts and their convergence into irreversible anatomical organ outcomes. Previous clocks have lacked histological definition because tissue sections are typically accessible only through invasive or postmortem sampling. Anatomical clocks derived from MRI or radiography appear intuitive but reflect delayed consequences that obscure progressive tissue degeneration. In contrast, molecular clocks based on peripheral blood rely on the assumption that organs release measurable signals into circulation.^12^ Although enrichment mapping has been used to infer origins, these attempts remain pseudo-specific, as signal abundance varies markedly across organs where weakly secretory organs such as the retina are easily overwhelmed by highly secretory ones such as the liver. Another concern is that transient molecular drifts do not necessarily correspond to histological degeneration or anatomical organ outcomes.^16,17^ While our approach is unique in enabling histological aging readouts *in vivo*, we expect the hierarchical aging framework may have similar impacts in other organs.

Neuroectodermal aging within the retina is a strong predictor of both mortality and longevity. Evolutionary theory posits aging as an acceleration of mortality risk; therefore, the validity of any aging clock should lie in its ability to calibrate this risk.^34^ Time-to-event analysis formalizes this by estimating the instantaneous hazard over time, within which CHRONOS robustly predicted mortality. Yet mortality captures only how rapidly homeostasis deteriorates, whereas longevity reflects the opposite end where resilience against entropic decline is preserved. The striking ability of CHRONOS to anticipate both mortality and longevity suggests that the neuroectodermal retina encodes life-course aging dynamics rather than being driven simply by the late-stage disease burden that often dominates mortality. While CHRONOS was trained on chronological age following standard conventions analogous to the first-generation clocks, we anticipate that incorporating mortality and longevity as ground truth endpoints may further improve its sensitivity to systemic deterioration, albeit at the cost of absolute age prediction accuracy.^11^

Aging assessment through the eye offers unique translational advantages. First, public engagement with eye health exceeds that for most organ systems, as individuals value vision above all other senses.^35^ This prioritization has translated into broad access to retinal imaging, with more than half of the UK population receiving regular eye examinations compared with only about one-tenth undergoing contemporaneous publicly funded cardiovascular screening.^36^ Second, the retina provides insights that extend beyond the eye. Our findings show that non-invasive retinal imaging can quantify aging and inform systemic health, a paradigm likely generalizable to other organ systems and rare conditions not represented here. Third, the accessibility of CHRONOS enables population-scale deployment. Unlike epigenetic and proteomic clocks that require invasive and costly assays, intracranial brain clocks that depend on MRI, or radiographic clocks that introduce ionizing radiation, retinal scanning is entirely risk-free, rapid, and readily integrated into routine care.^36^ We anticipate that a scalable eye clock will serve as both an organ-specific readout within multiorgan frameworks and a practical surrogate for systemic aging, bridging mechanistic insight with clinical translation. Continued validation across diverse populations and clinical settings remains warranted to accelerate its adoption in real-world healthcare.

Despite multiple strengths, this study has several limitations. First, CHRONOS was primarily developed in East Asian cohorts, which may limit its generalizability across ancestries. External validation in the UKB mitigated this concern, demonstrating cross-ethnic and cross-regional robustness of the captured neuroectodermal aging in the retina. While these patterns proved evolutionarily conserved across species, expanding recruitment to include more diverse populations from underrepresented regions is expected to further improve performance and representativeness. Second, the present retinal sections were acquired using an ∼850 nm spectral-domain system, and we expect a frequency-swept light source at longer wavelengths and deeper penetration would further improve performance. Yet, the steep signal attenuation at the melanin-rich RPE layer remains advantageous by confining sensitivity to neuroectodermal retina and minimizing contamination from the mesoderm-derived choroid. Third, although the prospective design and careful evaluation of reverse causation and confounding strengthen confidence in the observed associations, causal relationships remain to be confirmed through dedicated inference frameworks.

In conclusion, we demonstrate that optical biopsy of the retinal laminae coupled with deep learning enables real-time, *in vivo* profiling of neuroectodermal aging at histological granularity across primate species. The development of a neuroectodermal aging clock offers a robust framework for integrating tissue-level aging trajectories to health, mortality, and longevity, offering a tractable bridge between mechanistic insight and population-scale health surveillance.

## Methods

### Human cohort participants

Three healthy human cohorts (COIP, GTES, and ZAP) were enrolled to develop the CHRONOS clock, and five independent cohorts (SCOPE, UKB, LOVE, HOPE, and GDES) were used for external validation. Among the validation cohorts, SCOPE, LOVE, and HOPE comprised exclusively healthy participants, whereas the UKB represented a population-based cohort with mixed health status and the GDES comprised exclusively patients with diabetes. This study adhered to the tenets of the Declaration of Helsinki and were approved by the Ethics Committees of the Zhongshan Ophthalmic Centre, the London School of Hygiene and Tropical Medicine, and the Northwest Multicentre Research Ethics Committee. Written informed consent was obtained from all participants before enrolment.

Details of the COIP have been previously published.^37-39^ Briefly, the COIP is a bidirectional cohort that recruited over 15,000 healthy adults from Guangzhou, China, with both retrospective and prospective arms. Retinal tomography was performed during the baseline assessment of the prospective arm between 2017 and 2019 and was available for at least one visit in the retrospective arm. Retinal sections with signal strength <45, poor centration, or segmentation errors were excluded after manual inspection. Participants with eligible sections for both eyes were included for analysis (*n* = 12,579).

Details of the GTES project have been previously published.^40-42^ Briefly, the Guangzhou Twin Registry is a population-based registry of over 10,000 twin pairs residing in Guangzhou, China. Between 2006 and 2012, a subset of registry members was invited for annual ophthalmic examination including retinal tomography. The examined subset included (1) younger twin pairs (minors), whose parents were also invited and examined during the same visit, and (2) older twin pairs (adults). We restricted our analysis to adult participants only, which included the parents of the minor twins and the adult twins. All sections were manually inspected, and those with signal strength <45, poor centration, or segmentation errors were excluded. Adult participants with eligible sections for both eyes were included for analysis (*n* = 1,346).

Details of the ZAP study have been previously published.^43-45^ Briefly, the ZAP was a collaborative project among the Zhongshan Ophthalmic Centre, Moorfields Eye Hospital/University College London, and the Wilmer Eye Institute. The study screened over 10,000 participants and followed 889 primary angle-closure suspects (PACS) between 2008 and 2023, which demonstrated an extremely low risk of vision impairment and concluded that iridotomy for PACS was unnecessary. During the study, a subset of participants along with their accompanying family members or friends were invited for retinal tomography. The analysis was restricted to healthy participants with eligible sections for both eyes, where sections with a signal strength <45, poor centration, or segmentation errors were excluded (*n* = 867).

The SCOPE study is a prospective, multicentre study that recruited over 6,000 healthy participants across four assessment centres (three urban and one rural) in southern China between 2011 and 2014. Retinal tomography was performed at baseline in the three urban centres and introduced at follow-up in the rural centre (Yangxi) between 2020 and 2021. Sections with a signal strength <45, poor centration, or segmentation errors were excluded after manual inspection. Participants with eligible sections for both eyes were included for analysis (*n* = 4,918).

Details of the UKB study have been previously published.^12,18,46^ Briefly, the UKB is a prospective, multicentre study that recruited over 500,000 participants across the UK between 2006 and 2010. Retinal tomography was introduced in 2009 and performed during initial follow-up visits at 19 assessment centres. Fourteen centres with fewer than 100 participants were excluded. Consistent with UKB quality control procedures previously published, sections with signal strength <45 and within the lowest 20% for the inner limiting membrane indicator, validity count, or motion indicator were excluded. Participants with eligible sections for both eyes were included for analysis (*n* = 34,726).

Details of the GDES study have been previously published.^32,47,48^ Briefly, the GDES is an ongoing, prospective cohort study that recruited over 2,500 patients with type 2 diabetes in Guangzhou, China. Baseline assessments were conducted between 2017 and 2019 (*t*_0_), followed by biennial follow-ups in 2019–2021 (*t*_1_), 2021–2023 (*t*_2_), and continuing since 2023 (*t*_3_). Retinal tomography was performed at every visit, enabling longitudinal assessment of within-individual reproducibility and aging trajectories. All sections were manually inspected, and those with a signal strength <45, poor centration, or segmentation errors were excluded. Participants with eligible sections for both eyes at baseline were included for analysis (*n* = 2,041).

The LOVE study is an ongoing, prospective cohort study that screened over 10,000 adults with and without diabetes in Guangzhou, China, to investigate the longitudinal trajectory and metabolic determinants of optic neurodegeneration. All participants underwent retinal tomography at baseline between 2023 and 2024. Participants with signs of retinopathy or neuropathy, along with a subset of healthy individuals, were invited for an ongoing biennial follow-up over four years. The analysis was restricted to healthy participants with eligible sections for both eyes, excluding those with signal strength <45, poor centration, or segmentation errors (*n* = 8,235).

The HOPE study is an ongoing, prospective cohort study that recruited over 2,000 participants with or without myopia in Guangzhou, China, to delineate natural trajectories of ocular structure and function decline during aging. All participants underwent retinal tomography at baseline between 2022 and 2024 and were invited for an ongoing biennial follow-up over six years. The analysis was restricted to healthy participants with eligible sections for both eyes, excluding those with signal strength <45, poor centration, or segmentation errors (*n* = 1,890).

### Development of the neuroectodermal aging clock

We developed CHRONOS to estimate neuroectodermal aging within the retina. The system integrated a fully automated image preprocessing pipeline with a customized aging clock. The central tomographic sections of the retina from both eyes were identified and converted to greyscale. The sections were then spatially aligned by centring the fovea along the horizontal axis using a thickness-based curvature algorithm. Each section was rescaled to a 512 × 512-pixel matrix, and an artifact suppression procedure combining median and Gaussian filtering was applied to reduce speckle noise. To prevent data leakage, pixel intensities were normalized using parameters derived solely from the training data. Sections from both eyes were concatenated into a two-channel tensor. Small-angle rotation and elastic deformation were applied to improve model generalization.

The CHRONOS clock is a hybrid convolution–Transformer neural network designed to estimate chronological age among healthy individuals from retinal sections. To establish the reference age scale, chronological age was defined as the interval between the date of assessment and birth. The network integrates convolutional encoding with Transformer-based contextual reasoning to bridge fine-grained textures with long-range dependencies across the neuroectodermal retina. In the network front end, sequential residual convolutional blocks with batch normalization and rectified linear unit activations transform tomographic inputs into multiscale digital tissue representations. These layers condense local contrast and microtextural cues into hierarchically organized embeddings, while residual projections preserve information continuity across convolutional stages to ensure that fine tissue details remain accessible to deeper layers.

The Transformer encoder subsequently integrates these embeddings through multi-head self-attention, dynamically modelling spatial and semantic dependencies within the neuroectodermal retina’s laminated organization. This mechanism enables the network to capture global structural relationships beyond the receptive fields of convolution, approximating contextual reasoning across the neuroectodermal retina. Successive residual and feed-forward layers further refine these dependencies, organizing tokenized laminar features into unified contextual representations that reflect the anatomical hierarchy of the retina. Finally, the aggregated feature maps are passed through a multilayer perceptron (MLP) head to generate a continuous neuroectodermal aging estimate.

Model optimization was performed using an L1 loss function against chronological age. AdamW optimizer was employed at an initial learning rate of 1×10^−5^ and a weight decay of 5×10^−4^. A ReduceLROnPlateau scheduler was employed by adaptively reducing the learning rate by 20% after five consecutive epochs without improvement, with a minimum rate threshold of 1×10^−6^. Early stopping was applied when validation loss failed to decrease for five epochs, with a maximum of 50 epochs allowed per training cycle. Internal validation was performed through five-fold nested cross-validation on the holdout test sets.

To benchmark the architecture, we compared the convolutional–Transformer neural network against four alternative backbones (VGG-16, Inception-v3, ResNet-50, and ViT-Base/16) under identical optimization and training-validation splits. Each architecture was adapted to accept a two-channel tensor input, and its final fully connected layer was replaced with an MLP head to match the CHRONOS output configuration. All models were trained using the same learning rate schedule and early stopping criteria, ensuring that performance differences reflected architectural characteristics rather than optimization bias. The number of model parameters was also extracted and compared across architectures.

The CHRONOS system was applied to five independent cohorts for external validation. Model performance was quantified using the MAE and Pearson’s r between predicted and chronological age of each population. For the GDES, model performance was evaluated using baseline assessments. To adjust for chronological age, we defined NAG as the residual from regressing neuroectodermal age on chronological age. By definition, the resulting NAG is uncorrelated with chronological age. The rate of neuroectodermal aging was estimated as (*EAG*_*i*_ - *EAG*_0_)/T, where *EAG*_0_ represents the age gap at baseline, *EAG*_*i*_ the age gap at the *i*^th^ follow-up, and *T* the follow-up years.

### Follow-up for hallmark-related ARDs and mortality

UKB participants were linked to the UK National Health Service (NHS) Hospital Episode Statistics database for hospital admissions and to the NHS Central Registry for mortality. The NHS provides most of the healthcare in the UK, including inpatient and outpatient services, and record linkage was performed using the unique NHS identifier assigned to all residents. ARD outcomes were classified according to the International Classification of Diseases-10, with records available through Nov 2024. Person days for each participant was calculated from the date of baseline assessment to the date of disease onset, death, or end of follow-up, whichever occurred first.

Nine fundamental cellular hallmarks of aging were quantified indirectly through an individual’s vulnerability to hallmark-related ARDs, following a previously validated multistep framework.^49,50^ These comprised four primary (GI, TA, EA, LP) and five compensatory and integrative hallmarks (DNS, MD, CS, SCE, AIC). For each hallmark, 30 ARDs previously shown to be strongly associated with that process were included, encompassing both hallmark-specific and shared conditions, yielding a total of 83 distinct ARDs. The presence of a given hallmark was defined by the occurrence of one or more related ARDs in an individual.

The attribution of ARDs to each specific hallmark followed a validated pipeline.^49,50^ First, diseases with incidence rates that increase with chronological age were mapped to hallmarks by text mining of the human aging corpus. Next, for each hallmark, 30 ARDs most strongly linked to cellular aging were validated through: (1) identification of shared genetic signals in the GWAS catalogue; (2) enrichment of hallmark-related gene ontology terms among encoded proteins; (3) enrichment of hallmark-related biological processes in the proteins representing the top ARDs; and (4) analysis of disease co-occurrence networks within each hallmark. Detailed information on ARD attribution is provided in **Supplementary Table S2**.

For the GDES participants, follow-up data were available for nine ARDs spanning all nine aging hallmarks. These included coronary heart disease, hypertension, lipoprotein disorder, chronic kidney disease, gout, diabetic retinopathy, glaucoma, age-related macular degeneration, and cataract. The follow-up data were retrieved until Jun 2025.

### Frailty and systemic aging phenotypes

Marker sets of systemic aging were derived from Argentieri et al.^51^ and were measured using baseline blood samples, physical examinations, and structured questionnaires among UKB and GDES participants. Field identifiers for all blood biomarkers and measures of physical and cognitive function are summarized in **Supplementary Table S5**. Quality control and adjustment for technical variation in blood-based biomarkers followed previously described procedures.^52^ Self-rated health, walking pace, facial aging, feeling tired/lethargic, and insomnia were coded as ordinal categorical variables in order of increasing frequency or severity. Sleeping more than 10 hours per day was coded as a binary variable using the continuous self-reported sleep duration.

BMI was calculated as weight divided by height squared, and WHR as waist circumference divided by hip circumference. SBP and DBP were averaged across repeated automated readings. Forced expiratory volume in one second (FEV_1_) was normalized by height squared, and dominant-hand grip strength was divided by body weight to account for body size. Arterial stiffness index was calculated as the time between peaks of the pulse waveform at the finger divided by height. Heel bone mineral density was estimated using the quantitative ultrasound index of the calcaneus. Reaction time was recorded as the mean latency to first button press across all correct matching trials, and the fluid intelligence score represented the total number of correct answers to 13 reasoning questions.

The comprehensive frailty index was computed using a validated algorithm^53^ summarizing systemic deficits across 11 domains, including sensory, cranial, mental, infirmity, cardiometabolic, respiratory, musculoskeletal, cancer, pain, and gastrointestinal systems (**Supplementary Table S6**). Leukocyte telomere length was quantified as the ratio of telomere repeat copy number (T) to that of a single-copy gene (*S; HBB*, encoding β-haemoglobin).^54^ The resulting T:S ratio was log-transformed and standardized based on the distribution of all individuals with available measurements.

### Validation in rhesus macaques

All animal care and procedures conformed to institutional guidelines approved by the Animal Care and Use Committee of Zhongshan Ophthalmic Centre, in accordance with the Principles for the Ethical Treatment of Non-Human Primates. Six adult rhesus macaques (*Macaca mulatta*) were fasted overnight and anesthetized with isoflurane. They were positioned prone on a stereotaxic cart and immobilized with vecuronium bromide. Cardiac and respiratory rates, blood oxygen saturation, and electrocardiogram were continuously monitored by a veterinary technician throughout the procedure. Retinal tomography was conducted in a dark room following pupil dilation with phenylephrine chloride (2.5%) and tropicamide (1%). Venous blood was collected for the measurement of ALT, AST, albumin, creatinine, triglycerides, LDL-c, and HDL-c.

To enable lifespan-scaled comparisons between humans and rhesus macaques, we calibrated species-specific age scales using sexual maturity and maximum lifespan anchors. Data on maximum lifespan and age at sexual maturity were obtained from the AnAge project.^55^ The relative progression of macaque age on the human scale *s*_*H*_ was estimated as 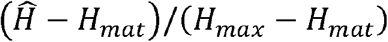, where *H*_*mat*_ and *H*_*max*_ denote human sexual maturity and maximum lifespan, respectively. The macaque-equivalent age was then estimated as *M*_*mat*_ + *s*_*H*_ × (*M*_*max*_ - *M*_*mat*_ ), where *M*_*mat*_ and *M*_*max*_ denote macaque sexual maturity and maximum lifespan, respectively. NAG for each macaque was derived as the deviance of the lifespan-rescaled CHRONOS age estimate and its chronological age.

### Mapping digital representations to retinal tissues

To characterize global feature representations, activations from the final encoder layer were extracted for all individuals. These activations constituted latent embeddings of the learned tissue architecture. Each feature map was flattened and projected into a two-dimensional manifold using UMAP. Data were stratified by cohort, including the aggregated training cohorts (COIP, GTES, ZAP) and testing cohorts (SCOPE, UKB, GDES), and randomly downsampled to a maximum of 5,000 datapoints per cohort for visualization. The resulting embeddings were displayed with colour gradients corresponding to chronological age.

Pixel-wise attribution was quantified using gradient-weighted activation mapping. Gradients of the model output with respect to feature maps from the final convolutional layer were backpropagated and spatially averaged to obtain channel-wise weights. These weights were linearly combined with the corresponding activation maps to generate saliency heatmaps highlighting structural vulnerabilities most predictive of neuroectodermal aging. The saliency maps were upsampled to match the original image resolution and smoothed with a Gaussian kernel to improve spatial coherence.

To evaluate causal contributions of anatomical substructures, we conducted counterfactual simulations using 1,000 manually delineated binary masks across six retinal laminae: RNFL, GC–IPL, INL, OPL–ONL, PL, and RPE. All masks were independently reviewed by a senior ophthalmologist (W.W.) to ensure boundary accuracy. Morphological operations were then applied to selectively perturb the geometry of each lamina while preserving the remaining architecture intact. Corresponding shifts in the predicted neuroectodermal age were quantified to assess the sensitivity of CHRONOS to laminar morphology.

To extend these analyses from individual sections to spatially resolved architecture, the three-dimensional laminar organization of the retina was reconstructed from consecutive tomographic sections. Laminar thickness was quantified using an automated segmentation algorithm based on graph search and multiscale gradient information.^56^ The method integrates local and global gradient cues to delineate layer boundaries with high spatial fidelity. Local edges were first detected using a Canny edge detector, while complementary global gradients were computed along the axial direction using a larger kernel. The two gradient sources were merged into a graph representation of the image, on which a shortest-path search via dynamic programming identified optimal layer boundaries. Mean thickness for the RNFL, GC–IPL, INL, OPL–ONL, PL, and RPE was subsequently derived for population-level association study.

### Statistical analysis

Data analyses were performed using R (v4.4.1) and Python (v3.10.12). Associations between NAG and aging biomarkers, physical/cognitive function measures, and thickness of retinal laminae were tested using multivariable linear or logistic models where applicable. Models were adjusted for chronological age, sex, ethnicity, assessment centre, Townsend deprivation index, physical activity group, and smoking status, as by Argentieri et al.^51^ As the GDES is a single-centre cohort with homogeneous ethnicity, adjustment for ethnicity and assessment centre was unnecessary. Unless otherwise specified, all *P* estimations were two-sided and corrected for multiple comparisons using the Benjamini-Hochberg method, with adjusted *P*⍰<0.05 considered statistically significant.

Associations for prevalent outcomes were estimated using multivariable logistic models, whereas incident outcomes were evaluated using multivariable Cox proportional hazards models. The latter modelled time from baseline to event onset, accounting for the right-censoring of the follow-up time. Fine–Gray hazards models were used to account for competing risks. The adjusted covariates were as described above, and multiple testing corrections were applied to all analyses. All prevalent cases were excluded from the corresponding incident analyses to ensure participants contributed only non-case follow-up time (e.g., in the case of GI, participants with all prevalent GI-related ARDs were excluded). Kaplan-Meier curves were generated at population-average covariate values within each relevant subject population.

In the longitudinal assessment of faster neuroectodermal aging, analyses were constrained in the GDES as it provides repeated assessments across timepoints. Associations involving the rate of NAG were modelled analogously to those for cross-sectional NAG but with additional adjustment for baseline NAG, ensuring that estimated effects were independent of both chronological age and baseline NAG. To test for accelerated neuroectodermal aging in diabetes and multimorbidity, the rate of NAG was regressed on the mean NAG across two timepoints. A positive association between these quantities indicated population-level acceleration, with the slope representing the estimated acceleration rate.

## Data Availability

All data produced in the present study are available upon reasonable request to the authors.

http://www.ukbiobank.ac.uk

## Data availability

UKB data are available upon reasonable request through the UKB data access procedures (http://www.ukbiobank.ac.uk). Permission to use the UKB resource was obtained via material transfer agreement under application No. 105658. GTES and ZAP data are available to qualified investigators upon request to principal investigators Mingguang He and Wei Wang. GDES, COIP, SCOPE, LOVE, and HOPE data are available to qualified investigators upon request to principal investigators Wei Wang. Data on animal lifespan and sexual maturity were obtained from the AnAge project (https://genomics.senescence.info).

## Code availability

All software used in this study is publicly available. The code used to develop the CHRONOS system is provided as a compressed .zip file during the peer-review process and will be made publicly available upon publication.

## Acknowledgements

This study was funded by the GBRCE for Major Blinding Eye Diseases Prevention and Treatment, the Hainan Province Clinical Medical Center, the National Natural Science Foundation of China (82371086), and the Science and Technology Projects in Guangzhou (SL2024A03J00472; 2025A04J7150). We thank all the participants and staff of the COIP, GTES, ZAP, SCOPE, UKB, LOVE, HOPE, and GDES.

## Author contributions statement

Study concept and design: W.W.; Acquisition, analyses, or interpretation: S.Y., Z.X., W.W.; Drafting of the manuscript: S.Y., Z.X., W.W.; Critical revision of the manuscript for important intellectual content: W.W.; Statistical analyses: S.Y., Z.X., W.W.; Obtained funding: W.W.; Administrative, technical, or material support: S.Y., W.W.; Study supervision: W.W.

